# Simulation-Based Comparison of Controlled Interrupted Time Series (CITS) and Multivariable Regression

**DOI:** 10.64898/2026.04.10.26350670

**Authors:** Fredrick Orwa, Charles Mutai, Ildephonse Nizeyimana, Ann Mwangi

## Abstract

When randomized controlled trials are impractical, interrupted time series designs offer a rigorous quasi-experimental approach to assess population level policies. Indeed, in the context of quasi-experimental designs (QEDs), the Interrupted Time Series (ITS) method is commonly thought of as the most robust. But interrupted time series designs are susceptible to serial correlation and confounding by time-varying factors associated with both the intervention and the outcome, which may result in biased inference. Thus, we provide a simulation-based contrast of controlled interrupted time series (CITS) and multivariable regression (multivariable negative binomial regression) for estimation of policy effects in count time series data. These approaches are widely used in policy evaluations, yet their comparative performance in typical population health settings has rarely been examined directly. We tested both approaches within a variety of data generating situations, differing in the series length, intervention effect size, and magnitude of lag-1 autocorrelation. Bias, standard error calibration, confidence interval coverage, mean squared error, and statistical power were assessed for performance. Both methods gave unbiased estimates for moderate and large intervention effects, although bias was more pronounced for small effects, particularly in short series. Although the point estimate performance was similar, inferential properties varied significantly. CITS always had smaller mean squared error, better consistency between model based and empirical standard errors, and confidence interval coverage near the 95% nominal levels over weak to moderate autocorrelation. By contrast, multivariable regression was more sensitive to serial dependence, leading to underestimated standard errors and undercoverage, especially at moderate to high autocorrelation, regardless of Newey-West adjustments. These findings show the benefits of using a concurrent control series and the importance of structurally accounting for serial correlation when studying population level policies with time series data.

## 1 Introduction

An interrupted time-series approach emerges as such a powerful quasi-experimental design, particularly when randomized controlled trial is impractical and when an intervention has been introduced at the population level [1, 2]. Indeed, among quasi-experimental designs, the Interrupted Time Series (ITS) approach is considered the most rigorous [3]. It gives a good analytical approach for evaluating the influence of policies and interventions in health, which were put in place at specific points in time [3]. It is of particular benefit in that it quantifies population-level intervention effects, which may encompass both direct and indirect (herd) effects [2]. ITS methods analyze aggregated data taken at regular time intervals before and after the intervention [4, 5]. The Interrupted Time Series (ITS) designs have gained recognition for methodological rigor, compared to traditional before and after studies, and are considered one of the most efficient methods for assessment of large-scale interventions and national-level policy interventions [3]. Although possessing many advantages, ITS analyses have certain methodological challenges. Therefore, some statistical methods have been proposed to study ITS data and address these methodological challenges. One widely regarded difficulty of ITS work is residuals in general are non-stationary or autocorrelated, so are not independent errors, with the possibility of providing erroneous inferences, usually via underestimated standard errors [6]. When the outcome is expressed as a count, an additional issue arises: the equality between the mean and the variance is assumed in the Poisson distribution, but health data tend to show greater variability than the mean, a condition known as overdispersion. In such cases, Poisson regression constrains the variance structure, leading to underestimated standard errors. To address this, negative binomial regression is preferred, as it introduces a dispersion parameter to accommodate extra-Poisson variation [7]. Another key limitation is that ITS estimates can be confounded by background trends that might not be directly related to the intervention [2]. For example, with the introduction of a free maternity policy, infant mortality might decrease, which might also denote concurrent benefits in healthcare systems. Thus, solely attributing the decline to the policy would be problematic. One method to handle confounding is using segmented regression, where an intervention is represented as a change in level and slope of the time series [5, 6]. The simplicity of this method in implementation and interpretation is why it is widely used. However, it assumes that the linear trend observed prior to the intervention would continue after the intervention, an assumption that is often difficult to justify [6]. Another option is the controlled interrupted time series (CITS) design, which uses a control series that is unaffected by the intervention but subject to roughly the same confounders before the policy [2, 3, 8]. Furthermore, multivariable regression remain in applied work, as in the assessment of the effect of ten-valent pneumococcal conjugate vaccine (PCV10) on invasive pneumococcal disease in Kenya [9]. These methods estimate policy effects using regression to adjust for measured confounders of the association between policy and outcome. Thus, in this study, we carry out a simulation-based comparison of controlled interrupted time series and multivariable regression methods for estimating policy effects subject to changing data generating conditions. More specifically, we evaluate their performance at various levels of lag-1 autocorrelation, time-series duration, and intervention effect size. These methods are generally common in policy evaluations, but their comparative performance in these common contexts has not been rigorously compared since they are used independently within the context of population healthcare interventions.

## 2 Materials and methods

### 2.1 Controlled interrupted time series (CITS)

Because we simulated a count outcome *Y*_*t*_ and a count control series *Z*_*t*_ that shared similar confounders in the pre-policy period, and imposed comparable confounder effects on both series, we fitted a negative binomial controlled interrupted time series (CITS) model with the control series included as an offset [2]. The model specification is given in equation 1.

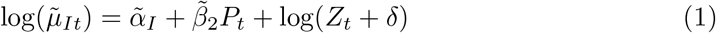

where 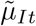 denotes the mean of the intervention series *Y*_*t*_, and δ is a small positive constant added to ensure that the logarithm of the control series is defined when the control series count is zero. 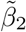 denotes the constant policy effect (level change) in our model. This specification is appropriate in our simulation setting because the data were generated under a constant policy effect scenario, with the intervention operating as a fixed level change over time.

#### 2.1.1 Motivation via log-ratios

To motivate the control-offset specification in equation (1), consider the (positive) control mean *µ*_*Zt*_ = 𝔼 (*Z*_*t*_) and intervention mean *µ*_*It*_ = 𝔼 (*Y*_*t*_). Under a common-trend assumption, both series share the same time-varying confounders (we assume also identical effects of the shared confounders), while only the intervention series is affected by the policy (in our case, both series share the temporal trend *t* and the confounder *X*_1,*t*_). On the log scale,

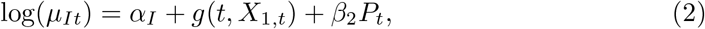

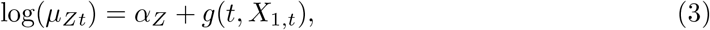

where *g*(*t, X*_1,*t*_) collects shared components (e.g., secular drift, seasonality, and other common confounding).

Subtracting (3) from (2) yields a log-ratio model:

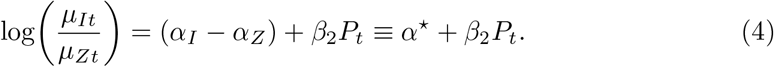

Therefore, under a constant level-change scenario,

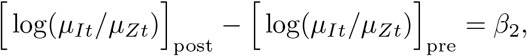

so *β*_2_ represents the policy effect on the log-IRR scale and exp(*β*_2_) is the corresponding incidence rate ratio (IRR).

From a causal perspective, this contrast compares the observed post-policy log-ratio to the counterfactual log-ratio that would have prevailed in the absence of the intervention, as proxied by the control series under the common-trend assumption.

Rearranging (4) gives

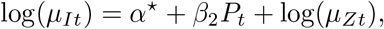

which motivates including the observed control series as an offset, i.e., 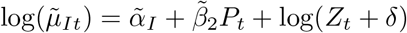 as in equation (1), where δ > 0 is included solely to accommodate occasional zero counts in *Z*_*t*_.

### 2.2 Multivariable negative binomial model

Consistent with the simulation design, we fitted a multivariable negative binomial regression model to the outcome (intervention) series *Y*_*t*_. The negative binomial specification was used to accommodate overdispersion introduced during data generation. The model is specified as

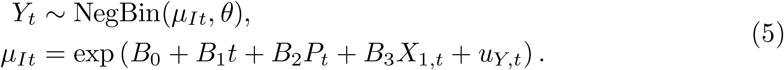

Here, *P*_*t*_ denotes the post-policy indicator, *t* represents time elapsed since the start of the study, and *X*_1,*t*_ is a time-varying covariate included to adjust for confounding as specified in the simulation framework. The coefficient *β*_0_ denotes the baseline level of *Y*_*t*_, *β*_1_ represents the coefficient of time (*t*), *β*_2_ indicate the constant policy effect in our case, and *β*_3_ corresponds to the effect of *X*_1,*t*_. Residual serial correlation was incorporated through an AR(1) process, *u*_*Y,t*_.

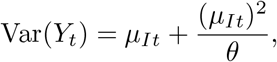

which allows the variance to exceed the mean.

#### 2.2.1 Interpretation of the policy effect in the multivariable model

To clarify the interpretation of the policy effect in the multivariable model, consider the expected outcome mean at the same time point *t* under the counterfactual pre-policy condition (*P*_*t*_ = 0) and the observed post-policy condition (*P*_*t*_ = 1). From equation (5),

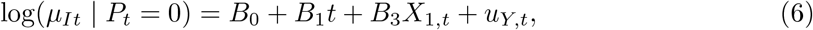

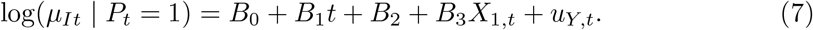

Subtracting (6) from (7) yields

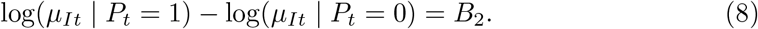

Thus, *B*_2_ represents the policy-induced level change on the log scale, comparing the observed post-policy outcome to the counterfactual outcome that would have occurred at the same time point in the absence of the intervention, conditional on the temporal trend and measured confounders. Exponentiating *B*_2_ gives the corresponding incidence rate ratio.

### 2.3 Simulation study methodology

We performed a numerical simulation study to compare the performance of controlled interrupted time series (CITS) and multivariable negative binomial regression models across a range of conditions. The simulated scenarios varied with respect to the presence of level change (constant immediate effect) in count outcomes, time-series length, and the degree of lag-1 autocorrelation. All design parameters were combined using a fully factorial design, with 8,688 simulated datasets generated for each combination.

The number of replications was calibrated based on the most restrictive scenario, defined by a short series length (*n* = 12), strong autocorrelation (*ρ* = 0.8), a small intervention effect (−0.0408), and the multivariable regression model. This scenario required the largest number of replications to ensure that the Monte Carlo standard error of the bias was below 0.005 and was therefore adopted for all other scenarios, for which the required number of replications was smaller. This calibration was informed by 100 pilot simulations (see S1 Table). Model performance was assessed using several predefined criteria. The simulation study was structured according to the ADEMP framework, encompassing the study aims, data-generating mechanisms, estimands, statistical methods, and performance measures [10].

#### 2.3.1 Data-generating process

We simulated a count outcome (intervention) series *Y*_*t*_ from a parametric negative binomial model with a single intervention introduced at the midpoint of the series and first-order autoregressive dependence of varying magnitude. Specifically, the outcome process was generated as in equation (5).

A control series *Z*_*t*_ was simulated in parallel using a similar parametric specification,

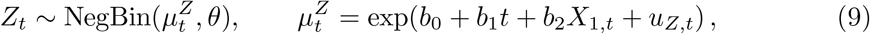

sharing the same magnitude of lag-1 autocorrelation, comparable temporal trends, and identical effects of the confounder *X*_1,*t*_, but with no direct policy effect. This construction ensured that the control series captured common underlying dynamics and confounding structures while remaining unaffected by the intervention.

The time-varying confounder *X*_1,*t*_ was generated to reflect a plausible policy-related shock combined with seasonal variation. Specifically, *X*_1,*t*_ was defined as

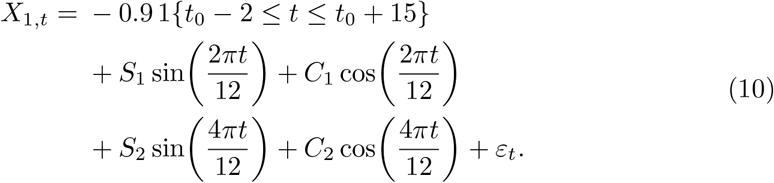

where 𝕀{·} denotes an indicator function capturing a transient shock around the intervention time *t*_0_, and *ε*_*t*_ ∼ 𝒩 (0, 0.2^2^). The seasonal amplitudes (*S*_1_, *C*_1_, *S*_2_, *C*_2_) were specified on the natural logarithmic scale.

Serial correlation in the outcome and control series was introduced through a first-order autoregressive process on the log-mean scale, with partially shared innovations between the intervention and control series. Specifically, we generated latent error processes for the outcome and control series as

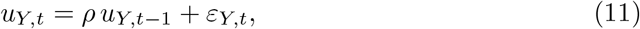

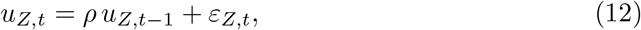

where (*ε*_*Y,t*_, *ε*_*Z,t*_) are bivariate normal innovations with mean zero and covariance matrix

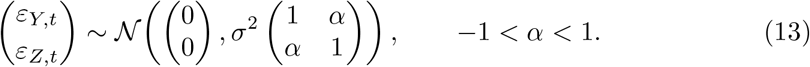

This construction implies Corr(*ε*_*Y,t*_, *ε*_*Z,t*_) = *α*, indicating that the two series share most, but not all, underlying shocks. In this study we set *α* = 0.85 to represent 85% correlation between the innovation shocks driving *Y*_*t*_ and *Z*_*t*_. When *α* = 1 the innovations are perfectly shared, whereas *α* = 0.85 produces strong but imperfect shared shocks, consistent with our simulation objective of producing an idealised control series while remaining realistic.

The process was initialized from its stationary distribution to ensure constant variance over time. The Simulation parameters for *Y*_*t*_,*Z*_*t*_, and confounders were informed by review of a few policy evaluation studies (S2 Table). A range of simulation scenarios was generated by systematically varying the intervention effect size (small, moderate, and large), the magnitude of lag-1 autocorrelation, and the length of the time series (Table 1). We did not consider positive intervention effect sizes, as the performance of the evaluated methods is not expected to depend on the direction (sign) of the policy effect.

**Table 1.** Summary of simulation parameters.

| Parameter | IRR (unexponentiated)/Any other | ln(IRR) |
| --- | --- | --- |
| Policy effect $B_2$ (Small effect) | 0.96 | -0.0408 |
| Policy effect $B_2$ (Moderate effect) | 0.70 | -0.3567 |
| Policy effect $B_2$ (Large effect) | 0.60 | -0.5108 |
| Outcome and control drift $B_1 = b_1$ (per time unit) | 0.995 | -0.0050 |
| Outcome intercept $B_0$ ($Y$; mean level 700) | — | $\ln(700) = 6.5511$ |
| Control intercept $b_0$ ($Z$; mean level 900) | — | $\ln(900) = 6.8024$ |
| Coefficient of $X_1$ ($B_3 = b_2$) | — | 0.1937 |
| AR(1) autocorrelation $\rho$ | 0, 0.20, 0.40, 0.60, 0.80 | — |
| Series length $n$ (time points) | 12, 14, 16, 18, 20, 24, 28, 32, 36, 40, 44, 48, 52, 56, 60, 80, 100, 150 | — |
| White-noise variance ($\sigma^2$ for both $\varepsilon_{Z,t}$ and $\varepsilon_{Y,t}$) | 0.10 | — |
| Corr($\varepsilon_{Y,t}, \varepsilon_{Z,t}$) | 0.85 | — |
| NB dispersion $\theta$ | 350 | — |
| Shock in $X_1$ (magnitude) | -0.9 | — |
| Seasonal amplitude $S_1$ | 1.04 | 0.0392 |
| Seasonal amplitude $C_1$ | 0.98 | -0.0202 |
| Seasonal amplitude $S_2$ | 1.04 | 0.0392 |
| Seasonal amplitude $C_2$ | 1.01 | 0.0100 |

#### 2.3.2 Estimands and other targets

The main estimand in this simulation study is the policy effect parameter *β*_2_, which captures the intervention-related level change and is defined as the coefficient associated with the policy indicator *P*_*t*_. This estimand was selected because it is a commonly reported effect measure in applied studies.

#### 2.3.3 Statistical Methods for Interrupted Time Series Analysis

We applied controlled interrupted time series (CITS) analysis, as specified in equation (1), and a multivariable negative binomial regression model, as defined in equation (5), to estimate the policy effect. Model parameters were estimated using maximum likelihood methods for all analyses. For both models, standard errors were adjusted using Newey–West estimators to account for autocorrelation, with the lag length selected as integer part of ⌊*n*^1*/*4^⌋, where *n* denotes the number of time points (series length) [2].

#### 2.3.4 Performance measures

Methods performance was assessed in terms of bias, empirical standard error, model-based standard error, 95% confidence interval coverage, mean squared error (MSE), and statistical power (S3 Table).

#### 2.3.5 Coding and execution

All simulations and analyses were implemented using reproducible code written in R. Data generation, model fitting, and performance evaluation were fully automated to ensure consistency across scenarios and replications. A random number seed was set once at the beginning of the script to allow exact reproducibility of all results.

#### 2.3.6 Analysis of the simulated datasets

Data analysis and visualization were implemented in R (version 4.4.1). Graphical and tabular displays were used to summarize and compare the performance of the statistical methods across simulation scenarios.

## 3 Results

### 3.1 Bias of estimated intervention effect

Estimates from both methods were generally unbiased across most scenarios, especially for moderate and large effects. Bias was generally small for moderate and large effects across all autocorrelation levels and was typically less than 10% for both methods. Bias was more pronounced for small intervention effects, where percentage bias was more variable, especially at short series lengths. In these settings, the effect of autocorrelation was more evident, with high autocorrelation often associated with bias greater than 20% across both methods. However, bias still showed improvement as series length increased. For weak to moderate autocorrelation (*ρ* ≤0.4), both methods exhibited only mild bias once the series length reached approximately 40–50 time points for small effect sizes. Conversely, strong autocorrelation (*ρ* = 0.8) was associated with increased bias at short series lengths for small effects, and this bias appeared stronger for multivariable regression than for CITS.

The sensitivity analysis was conducted with the lag-1 autocorrelation of the control series fixed at 0.4, while the correlation between the shared shocks of the control and intervention series remained set at 0.85. The results closely resembled those from scenarios in which the control and intervention series shared the same magnitude of lag-1 autocorrelation, with only a slight reduction in precision. This indicates that the accuracy of estimates obtained from the CITS model is driven primarily by shared pre-policy confounders and common unobserved shocks, rather than by an exact match in the magnitude of autocorrelation between the intervention and control series (Fig 1, S1 Fig, and https://fredshiny.shinyapps.io/CITS_MULTIVARIABLE/).

**Fig 1.**
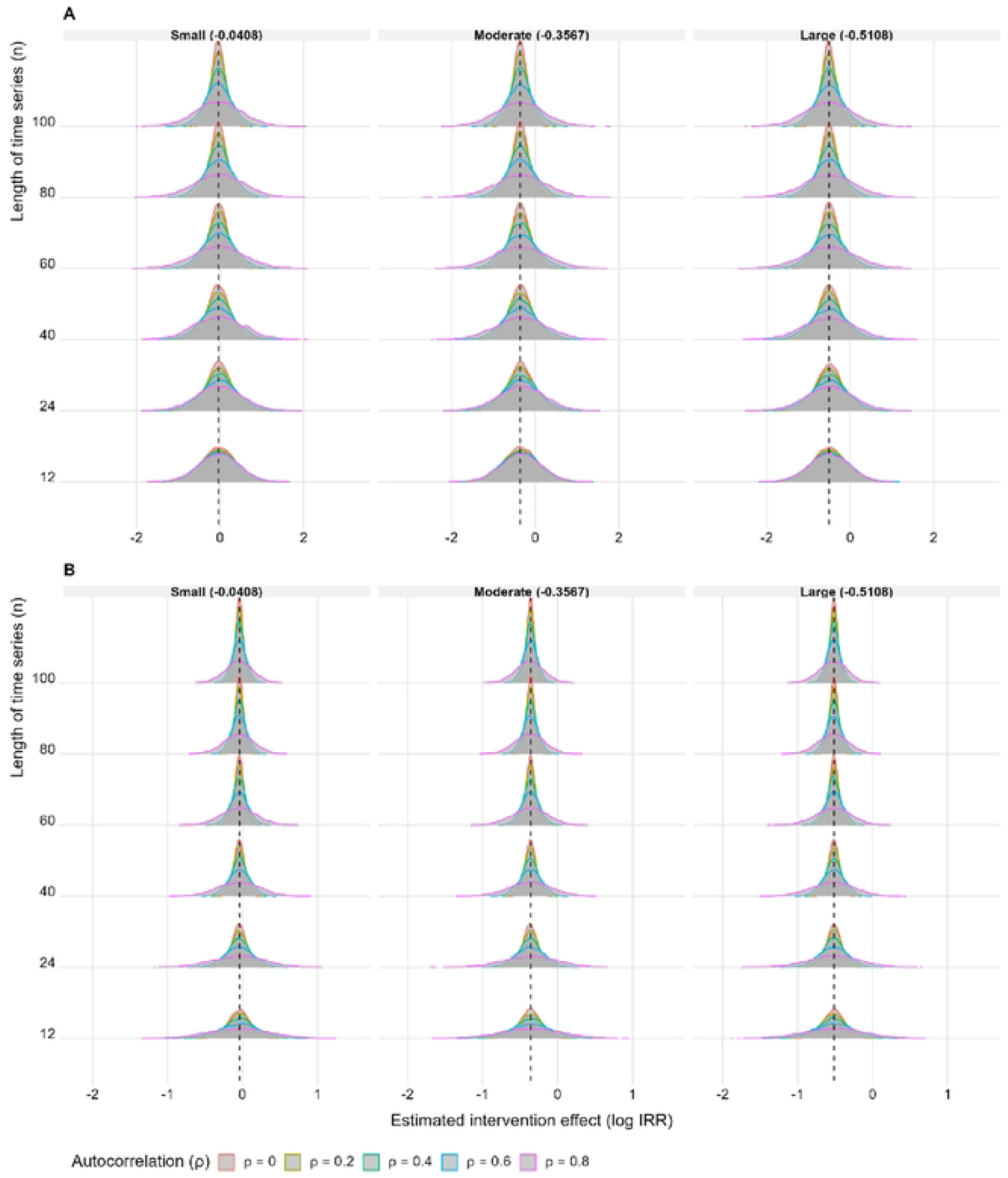
Distribution of policy effect estimates across scenarios. (A) Multivariable negative binomial regression. (B) Negative binomial model with control series included as an offset. The dashed vertical line denotes the true policy effect (estimand).

### 3.2 Empirical and Model-Based Standard Errors

Across both methods, empirical standard errors decreased with increasing series length, reflecting improved precision as more observations became available, with the strongest gains observed under weak autocorrelation (*ρ* ≤ 0.4).For both approaches, empirical and model-based standard errors increased with increasing autocorrelation, with a more pronounced increase observed for the multivariable regression than for CITS. In the multivariable regression, empirical standard errors increasingly exceeded model-based estimates as autocorrelation strengthened (*ρ* ≥0.6), indicating underestimation of uncertainty. In contrast, the CITS model exhibited smaller and better-aligned empirical and model-based standard errors across all scenarios, including under strong autocorrelation (*ρ* = 0.8) when series length was large, demonstrating more reliable uncertainty quantification. Across all scenarios, CITS exhibits smaller empirical standard errors than the multivariable negative binomial model, reflecting superior precision (Fig 2,Fig 3 and https://fredshiny.shinyapps.io/CITS_MULTIVARIABLE/).

**Fig 2.**
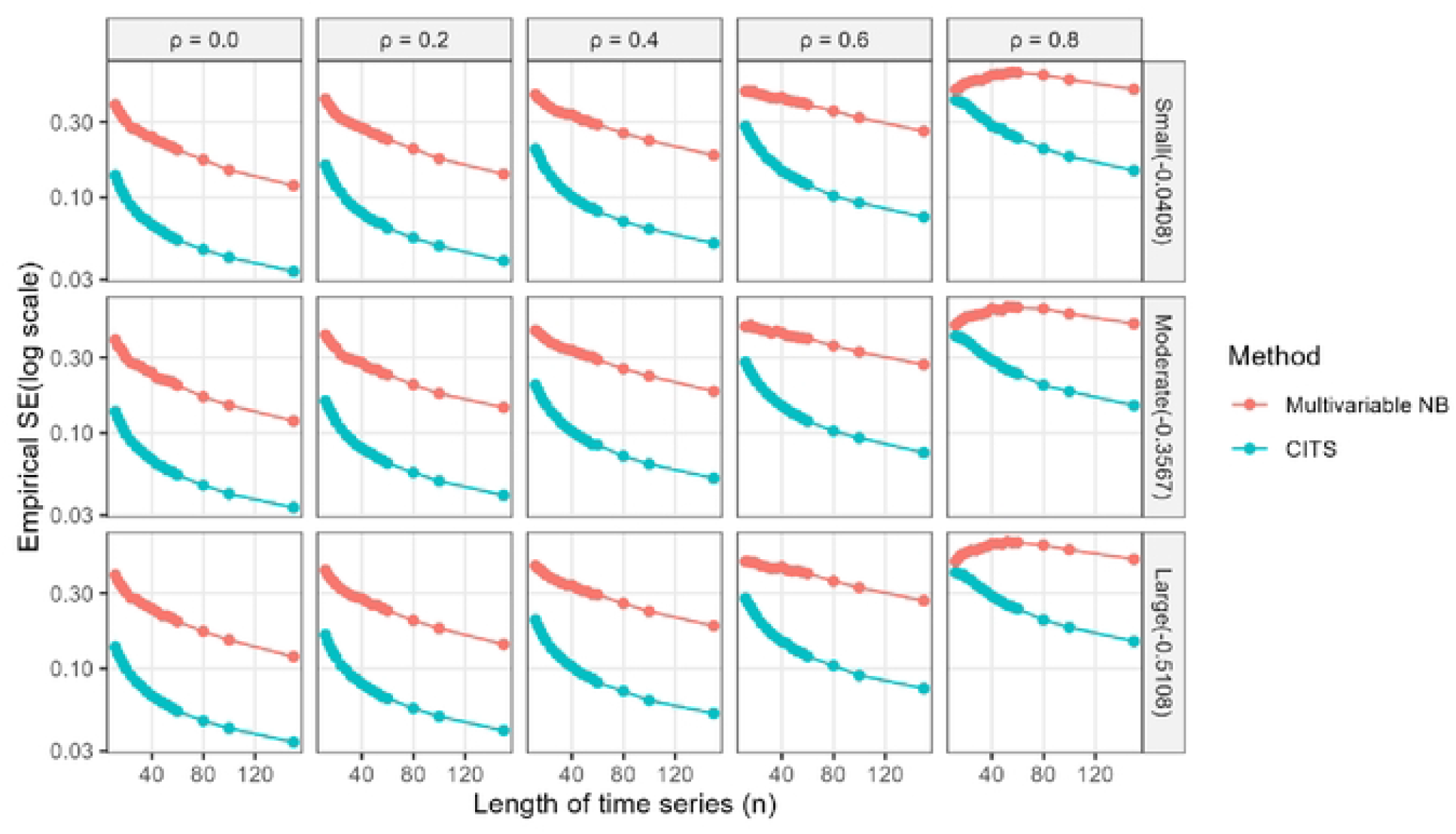
Empirical standard error comparison across effect size, autocorrelation levels, and series lengths.

**Fig 3.**
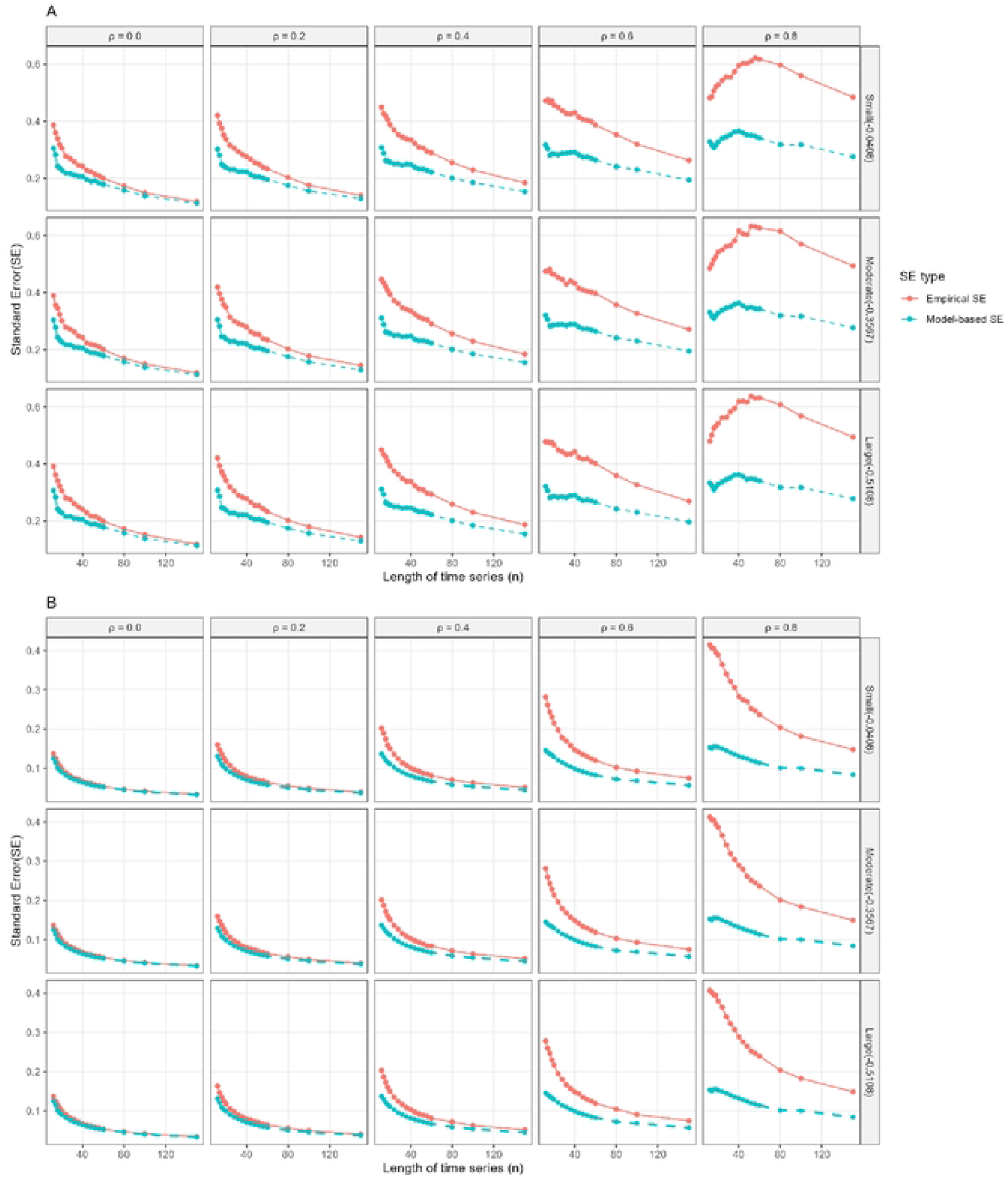
Standard error performance across scenarios. (A) Multivariable negative binomial regression. (B) Negative binomial model with the control series included as an offset (CITS).

In the sensitivity analysis, fixing the control series’ lag-1 autocorrelation at *ρ* = 0.4 had little impact on the empirical standard errors of the CITS estimator, even as outcome autocorrelation increased up to *ρ* = 0.8. Empirical standard errors remained stable and decreased smoothly with increasing series length, closely matching results from scenarios in which the control and outcome series shared the same magnitude of lag-1 autocorrelation. This suggests that reliable uncertainty quantification in CITS is driven primarily by shared covariates and common unobserved shocks, rather than by exact matching of serial dependence between the outcome and control series (S2 Fig and https://fredshiny.shinyapps.io/CITS_MULTIVARIABLE/).

### 3.3 Confidence interval coverage

The overall coverage of the nominal 95% confidence intervals varied for both methods. Coverage was generally below the nominal level for the multivariable regression and dropped rapidly with increasing autocorrelation. Coverage improved as the series length increased but remained well below 95% under moderate to strong autocorrelation (*ρ*≥ 0.4), even for large effect sizes.

By comparison, CITS provided better coverage in most cases, except in the presence of strong autocorrelation (*ρ* = 0.8). CITS achieved coverage close to the nominal level at moderate series lengths when autocorrelation was weak to moderate (*ρ*≤ 0.4), and coverage improved further as *n* increased. When autocorrelation was strong (*ρ* = 0.8), coverage fell substantially below the nominal 95% level and mirrored that of multivariable regression. Overall, CITS coverage was close to the nominal level, while multivariable regression suffered from obvious undercoverage, indicating substantial underestimation of uncertainty (Fig 4 and https://fredshiny.shinyapps.io/CITS_MULTIVARIABLE/).

**Fig 4.**
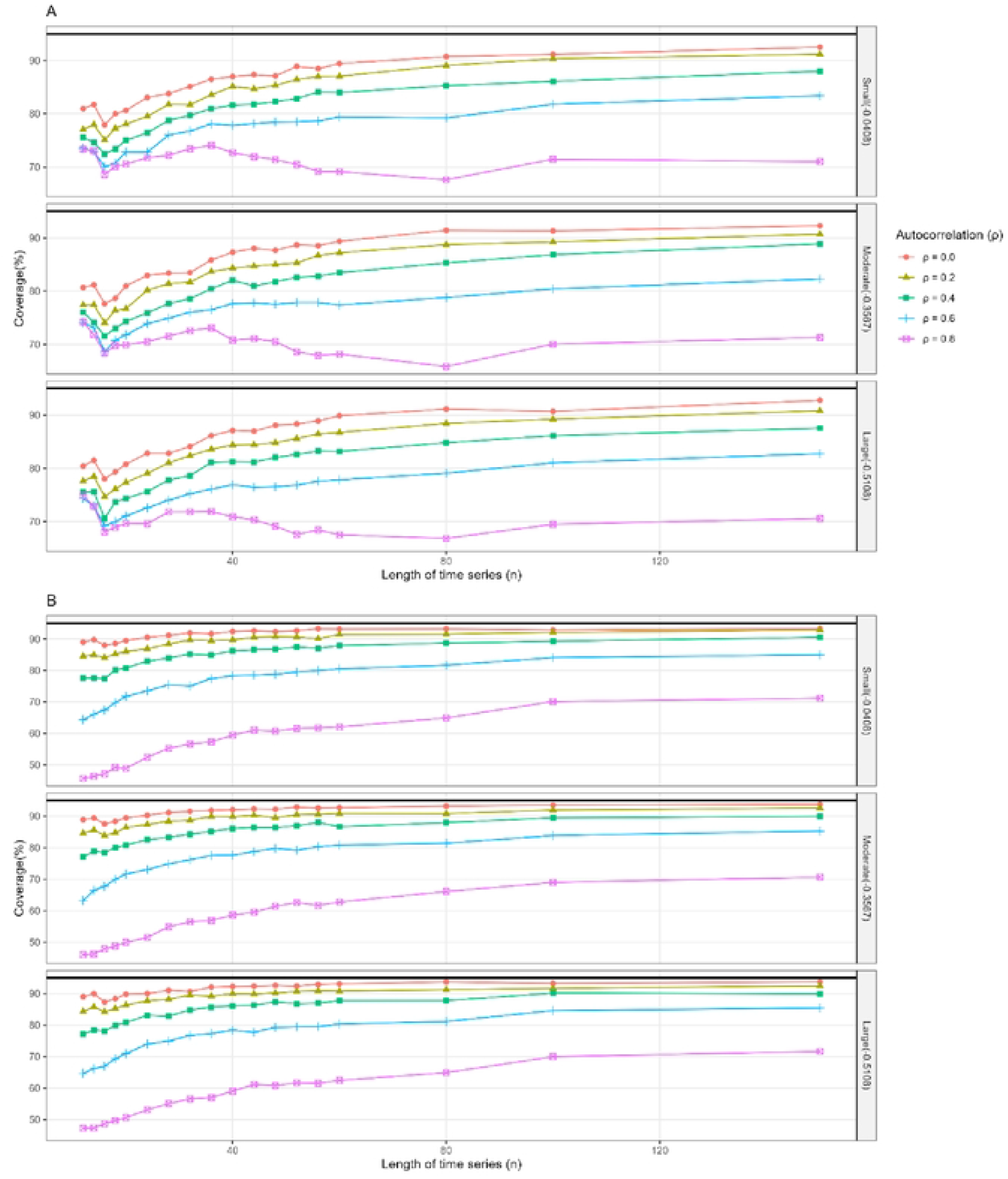
Coverage performance across scenarios. (A) Multivariable negative binomial regression. (B) Negative binomial model with the control series included as an offset (CITS).

### 3.4 Power

Power was interpreted not separately, but consistently alongside confidence-interval coverage. This is because power estimates can be misleading when standard errors are improperly calibrated and coverage does not attain the nominal 95%. In particular, scenarios with substantial undercoverage were not considered to demonstrate reliable detection behaviour.

Across all effect sizes, multivariable regression generally exhibited weak power, except under weak autocorrelation (*ρ* ≤ 0.2) with long series (*n* ≥ 80) and a large effect size, or under no autocorrelation (*ρ* = 0) with very long series (*n* ≥ 150) and a moderate effect size. Overall, power for the multivariable regression model was highly sensitive to increasing autocorrelation. Under a small intervention effect, power was low across all combinations of series length and autocorrelation (below 34%), indicating dependence of power on the size of the policy effect. For moderate and large effects, power increased slowly with increasing series length when autocorrelation was weak, but in most settings did not exceed the conventional 80% threshold. Crucially, these modest power levels coincided with substantial undercoverage, implying that the apparent power may not reflect well-calibrated inference. Multivariable regression therefore demonstrated poor and unreliable detection performance in the presence of serial correlation.

Similarly, power patterns for CITS closely mirrored the corresponding coverage results. When there was zero to moderate autocorrelation (*ρ* ≤0.4) and coverage was close to nominal, power increased sharply with series length for moderate and large effect sizes, approaching 100% in many scenarios. This indicates more accurate policy estimation with the inclusion of a control series, and the resulting power is trustworthy because coverage is near nominal. As would be expected, power was low for small effects (between 13% and 55%). Power was high under strong autocorrelation (*ρ* = 0.8), but this may be misleading, as it coincided with substantial undercoverage.

The results indicate that CITS has higher power to detect true effects when coverage is near nominal compared with multivariable regression, and that both methods have low detection power for very small intervention effects (Fig 5 and https://fredshiny.shinyapps.io/CITS_MULTIVARIABLE/).

**Fig 5.**
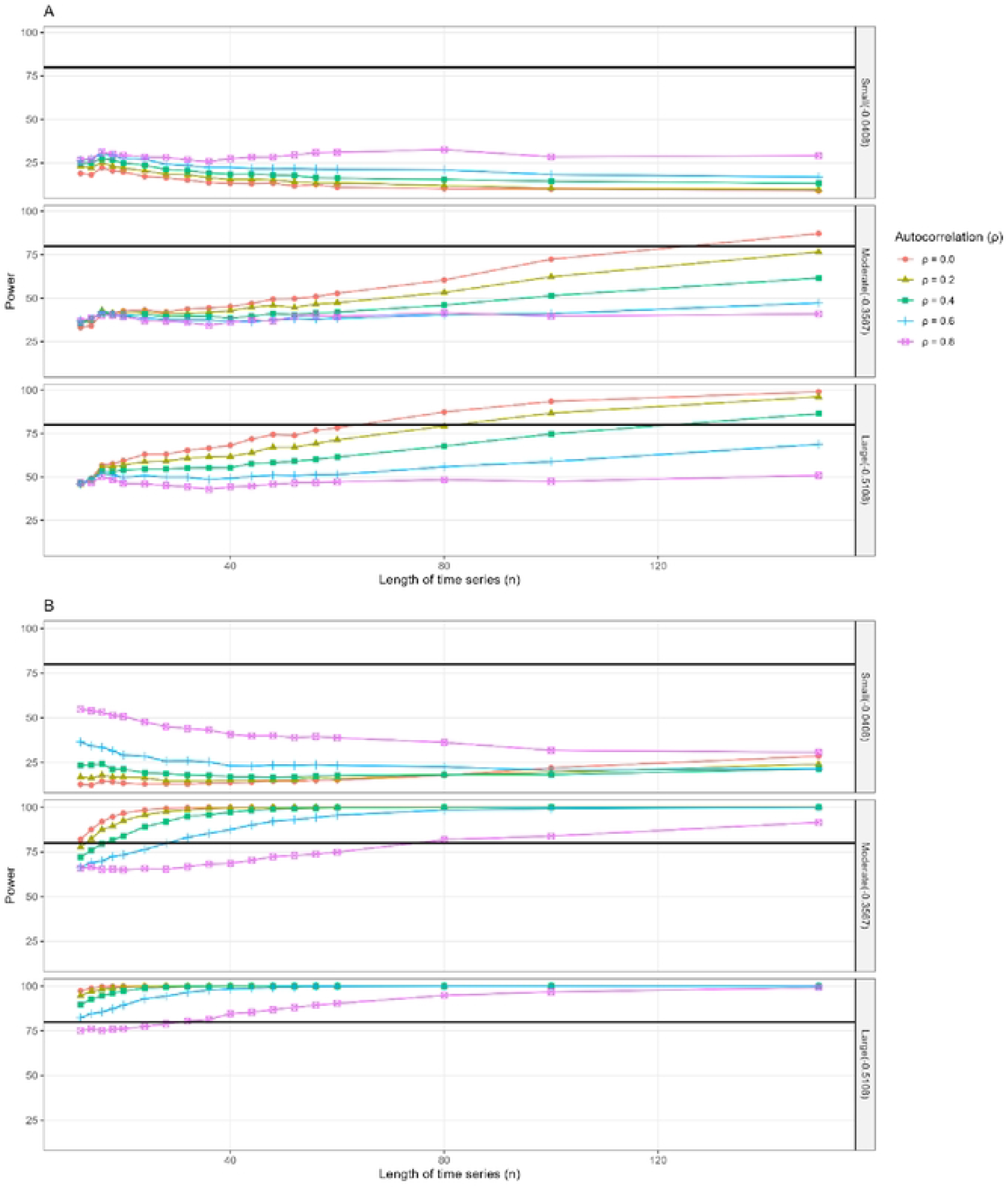
Power performance across scenarios. (A) Multivariable negative binomial regression. (B) Negative binomial model with the control series included as an offset (CITS).

### 3.5 mean squared error (MSE)

Fig 6 presents the mean squared error (MSE) of the estimated intervention effect across different levels of autocorrelation, series lengths, and effect sizes. The multivariable regression demonstrated larger MSE across all scenarios, with pronounced sensitivity to serial correlation. In particular, MSE increased rapidly as autocorrelation strengthened and remained elevated even for long time series under moderate to strong autocorrelation (*ρ* ≥0.4). Although increasing series length reduced MSE, the rate of improvement was slow under moderate to strong autocorrelation, suggesting persistent bias and variance inflation. Differences across effect sizes were small in comparison with the impact of autocorrelation.

**Fig 6.**
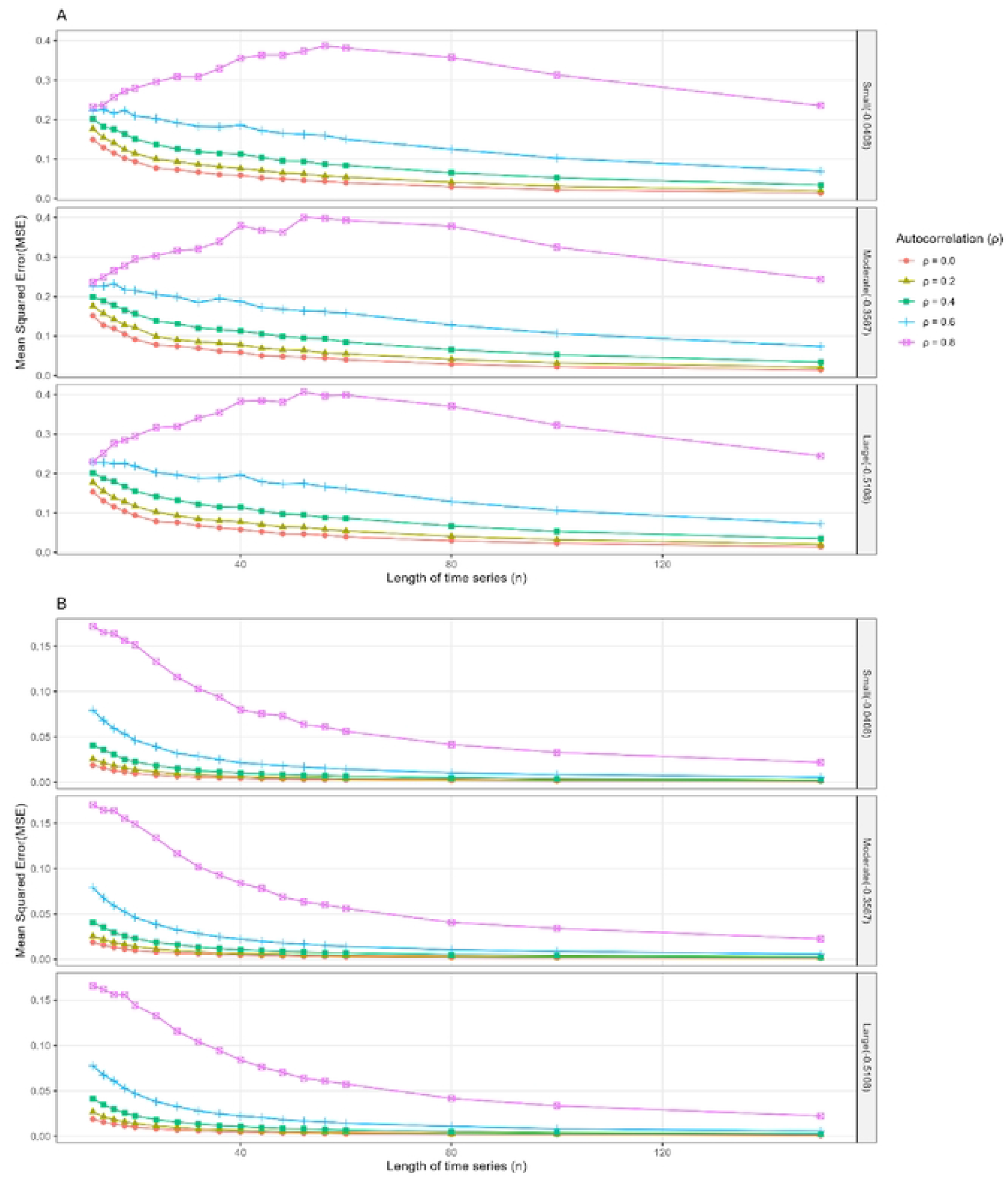
Mean squared error (MSE) performance across scenarios. (A) Multivariable negative binomial regression. (B) Negative binomial model with the control series included as an offset (CITS).

In contrast, the controlled interrupted time-series (CITS) approach achieved uniformly lower MSE across all scenarios. Even in the presence of strong autocorrelation (*ρ* = 0.8), MSE declined steadily with increasing series length and remained an order of magnitude smaller than that of the multivariable regression. For weak to moderate autocorrelation (*ρ* ≤0.4), MSE approached zero rapidly as *n* increased, indicating better bias control and improved estimation efficiency. These results demonstrate that CITS provides substantially more accurate effect estimation than multivariable regression in the presence of serial dependence. The specific estimates of MSE for both methods are also available at https://fredshiny.shinyapps.io/CITS_MULTIVARIABLE/.

## 4 Discussion

This study contributes to the expanding methodological literature investigating the performance of interrupted time series methods for intervention assessment, supplementing previous tutorials, review, and comparative studies [2, 6, 11]. Our simulation analysis results highlight differences in the performance of controlled interrupted time series and multivariable regression methods used to estimate policy impact under different scenarios, including differing effect sizes, series lengths, and degrees of lag-1 serial correlation. We chose these methods because they are widely used in practice, despite limited comparative evidence regarding their performance [9, 12].

Across the simulated scenarios, both controlled interrupted time series and multivariable regression resulted in largely unbiased estimates of the policy effect, except for small policy effects where bias seemed to be more pronounced. Bias for moderate and large effects, however, declined rapidly to relatively small percentages (less than 10%). Consistent with the literature, both methods remained approximately unbiased under autocorrelation [13]; however, moderate to strong autocorrelation slightly amplified bias, especially for both approaches in shorter series, particularly when the effect size was small. The CITS model showed closer agreement between model-based and empirical standard errors and also generally smaller empirical standard errors compared to multivariable regression across all scenarios, indicating greater precision. As previously presented in [2], the offset-based log-linear formulation cancels much of the shared serial dependence between the outcome and control series, whereas multivariable regression retains this dependence and may not fully account for it through Newey–West adjustment. Both methods generally produced coverage below the nominal 95% level. However, CITS achieved coverage closer to the nominal level when autocorrelation was low to moderate (*ρ* ≤0.4). These findings are consistent with the general principle described in [10], suggesting that undercoverage is expected when bias is non-zero and model-based standard errors underestimate empirical variability. In both the CITS model and multivariable regression, power increased with increasing series length and larger effect sizes. Power appeared to drop substantially with increasing autocorrelation in multivariable regression, most clearly for moderate and large effects; however, this apparent power was also associated with serious undercoverage. In contrast, the CITS model exhibited relatively stable power across autocorrelation levels when effect size was moderate to large. This pattern is consistent with findings reported in [14, 15].

Our study is consistent with the MSE patterns observed in prior simulation evidence showing that serial dependence substantially inflates estimation error and that increasing series length alone does not fully mitigate this effect [13]. Compared to regression-based methods assessed in prior work, the offset-based CITS formulation further reduces MSE by structurally cancelling the shared dependence between the outcome and control series [13], and its MSE was smaller than that of multivariable regression, especially when autocorrelation was moderate to large.

## 5 Conclusion

We have performed a simulation comparing a controlled interrupted time series and standard multivariable regression for policy evaluation under realistic data generation conditions. For a wide range of series lengths and levels of serial correlation, both approaches provided largely unbiased estimates of policy effects for moderate and large effect sizes, while bias was more pronounced for small effects. As a result, inference should be exercised cautiously if the size of the anticipated policy effect is very small. That said, their inferential performance differed substantially. The controlled interrupted time series model reliably performed better than multivariable regression and provided consistently smaller mean squared error and better agreement between model-based and empirical standard errors, as well as confidence interval coverage close to nominal levels in cases of weak to moderate autocorrelation. In these same settings, CITS also demonstrated higher statistical power for moderate and large effects as series length increased. Multivariable regression, on the other hand, showed greater sensitivity to serial dependence, higher estimation error, and poorer uncertainty quantification, resulting in low statistical power for series lengths below 80, despite the use of Newey–West standard error adjustments. These results emphasize the benefits of including a concurrent control series when using time-series data to assess population-level policies.

## Data Availability

The minimal dataset and accompanying code required to reproduce the findings of this study are available in the Harvard Dataverse repository (https://doi.org/10.7910/DVN/ARRLTM).

## Supporting information

**S1 Table. Calculated formulas for performance measures**. The number of simulations was determined using a pilot run of 100 simulations under the most restrictive setting (n = 12, *ρ* = 0.8, small effect size = -0.0408), such that the Monte Carlo standard errors for bias, mean squared error, and both model-based and empirical standard errors were below 0.005, and those for coverage and power were below 0.5 percentage points.

**S2 Table. Summary of key studies informing parameter selection for the simulation**.

**S3 Table. Definitions of performance measures used in the simulation study**.

**S1 Fig. Results from the multivariable negative binomial regression (A), the controlled interrupted time series model (B), and the controlled interrupted time series model with control-series autocorrelation fixed at 0.4 (C)**.

**S2 Fig. Empirical standard errors from the multivariable negative binomial regression, the controlled interrupted time series (CITS) model in which the control series shares the same lag-1 autocorrelation magnitude (***ρ***) as the outcome series (***Y*_*t*_**), and the CITS model with autocorrelation fixed at** *ρ* = 0.4.

## Acknowledgments

The author was awarded a Sub-Saharan Advanced Consortium for Advanced Biostatistics (SSACAB) fellowship funded by the Science for Africa Foundation to [Del-22-009], with support from the Wellcome Trust and the UK Foreign, Commonwealth & Development Office.

The author would like to thank professor Christian Bottomley of the London School of Hygiene & Tropical Medicine for his mentorship at the KEMRI–Wellcome Trust and for his work, which forms the basis of this study.

## Notes

### Competing Interest Statement

The authors have declared no competing interest.

### Funding Statement

Yes

### Author Declarations

The research was approved by Moi University

